# Effects of Health Education to Reduce Stroke Recurrence by Controlling Modifiable Factors: A Randomized Controlled Trial

**DOI:** 10.1101/2024.10.16.24315632

**Authors:** Mahabuba Afrin, Sharif Uddin Khan, Subir Chandra Das, K. A. T. M Ehsanul Huq, Mohammad Shah Jahirul Hoque Chowdhury, Yasuko Fukuoka, Yasuko Fukushima, Michiko Moriyama

## Abstract

**BACKGROUND:** Health education could be an effective measure to prevent stroke recurrence; however, the evidence is still ambiguous. The aim of this study was to evaluate the effectiveness of a health education program among stroke patients and their family caregivers to reduce stroke recurrence.

**METHODS:** A parallel (1:1), open-label, prospective randomized controlled trial was conducted at the National Institute of Neuroscience & Hospital in Bangladesh between October 2022 and March 2024. We enrolled stroke patients who were discharged from the inpatient department or attended as an outpatient of the hospital. The primary endpoint was the recurrence rate of stroke within 12 months of intervention. The secondary endpoints were the rate of all adverse events related to stroke that occurred within 12 months of follow-up visits.

**RESULTS:** We analyzed data from 432 patients (216 in each group). We found recurrence was 14 (6.5%) in the IG and 8 (3.7%) in the CG after 28 days of first-stroke attack, and the difference was not significant (p=0.189). We observed stroke-related adverse events 116 (26.9%) (25.0% in IG and 28.7% in CG, p=0.294) including death 95 (22.0%) (18.1% in IG and 25.9% in CG, p=0.041) during 12-month period. Even though stroke patient’s medication adherence was significantly improved at 6-month and 12-month after intervention in IG (p<0.001 and p=0.002, respectively), all events and all causes of death were not significant after adjusted by the severity of stroke (mRS) (p=0.933, p=0.341, respectively).

**CONCLUSIONS:** The findings of this newly approached intervention can be used as the most reliable evidence for epidemiological figures for Bangladesh. Health education could reduce death and enhance medication adherence significantly among stroke patients. This study results suggest the importance of structuring the acute care system and health education programs to integrate into the health system for the management of stroke. However, an accurate diagnostic and follow-up system is crucial.

**CLINICAL TRIAL REGISTRATION:** https://clinicaltrials.gov/ct2/show/NCT05520034.

## Introduction

Stroke burden is a significant and growing public health concern. It is the second most fatal and third most leading cause of disability worldwide. Its prevalence is rising quickly in low- and middle-income countries.^1^ Every year, more than 13.7 million strokes take place globally, which leads to 5.5 million deaths. Although strokes are more likely to affect elderly individuals, there is an increased frequency of strokes among younger people.^2,3^ In 2019, the foremost risk factors for stroke were elevated systolic blood pressure, high body mass index, elevated fasting plasma glucose concentrations, and smoking.^4^ It is indicated that the risk of recurrent stroke is the highest within first few months after the initial event, and up to 25% of stroke survivors experience a recurrent stroke within five years.^5^ Recurrent strokes are associated with fatal outcomes, increased disability, and higher mortality rates compared to first-time strokes. Addressing modifiable risk factors is crucial for reducing the risk of stroke recurrence and improving long-term outcomes for stroke survivors.^5^

In Bangladesh, stroke was the top cause of death per 100,000 population for both males and females and all ages in 2019.^6^ In 2022, Bangladesh had a stroke rate of 79.9%, 15.7%, and 4.6%, which was caused by strokes that involved ischemic, hemorrhagic, and subarachnoid hemorrhage respectively, and affected 11.39 thousand people.^7^ One study conducted in a tertiary Medical University Hospital in Bangladesh evaluated the rate of recurrence of stroke and found that the cumulative recurrence rate was 14.7% at three months, 15.3% at six months, 17.3% at ninth months, and 20% at one year. The most frequent age group was > 75 years which represented 44.4% of recurrent stroke.^8^

Despite the advancement of thrombolytic therapy and acute stroke management, preventing the recurrence of stroke remains a significant challenge. Moreover, the ability of patients to manage complications and avoid underlying disorders like hypertension, diabetes, dyslipidemia, etc. are impacted by substandard primary care and a lack of referral mechanisms. Patients in underdeveloped healthcare systems are not consistently diagnosed or treated because of a lack of information, infrastructure, resources, shortage of equipment, diagnostic accuracy, lack of health education, and budgetary constraints, which prevent them from getting health checkups.^9,10^

One systematic review concluded that health education may improve blood pressure targets, but not improve other risk factors or reduce recurrent events, and patient education alone could not prevent the risk of recurrent stroke.^11^ A study in Bangladesh found that a small sample size of 150 was insufficient for identifying stroke recurrence frequency. A larger sample size and health education efforts were recommended to reduce recurrence.^8^ Another study involved a 321-sample size in Japan provided self-management health education for six months. This study reduced recurrence rate by half after the intervention, however, the difference was not statistically significant for the prevention of stroke recurrence.^12^

Since there is no established patient follow-up system and health education offered across the hospitals in many developing countries, this is the first intervention study to incorporate patient health education following discharge from acute care hospitals. Even hospitals provide health education at the time of discharge to improve patients’ self-motivation at home, stroke patients are found to lack of knowledge to follow medical advice.^13^ As most of the patients are taken care of by the family caregivers, therefore, their health education can significantly reduce risk factors and complications related to stroke.^14^

In Bangladesh, stroke patients’ health education regarding self-management skills at home from the hospital level is limited. The considerable reasons could be socio-cultural, economic, and political as well as poor planning and improper implementation of health policies and programs within the health system. Another study conducted in Bangladesh recommended that risk factors for recurrent stroke should be identified early, and interventions should be taken immediately to control them to prevent recurrence. Stroke patients and their family caregivers should know the importance of controlling the modifiable risk factors.^8^ Therefore, it is crucial to establish a well-organized patient-disease management education system.

Based on the above study findings, we hypothesized that health education programs among first-stroke patients and their family caregivers could reduce the stroke recurrence rate by controlling the modifiable risk factors. We tried to evaluate the effectiveness of health education programs in improving risk factors and reducing the recurrence of stroke and other stroke-related adverse events.

## Methods

### Study Design and Participants

This was a parallel (1:1), open-label, prospective randomized controlled trial conducted at the National Institute of Neuroscience & Hospital (NINS&H) in Dhaka, Bangladesh between October 2022 and March 2024. We enrolled participants who had their first stroke and were discharged after getting inpatient treatment or visited at the outpatient department of NINS&H, age 18 years and above, both sexes, and willing to participate in this study. Stroke included ischemic or hemorrhagic with subtypes that were diagnosed by a physician. A family caregiver, living with the patient, and provided support to them were also included. In Bangladesh, ischemic stroke accounts for 79.9% of all stroke cases,^7^ and health education can be effective for patients with ischemic stroke classified as modified Rankin Scale (mRS) scores of 0-2. Considering the impact of self-management for hemorrhagic stroke, patients with an mRS score below 3 were included. As most of the stroke patients in Bangladesh have their family caregivers to provide healthcare support, in considering family education, patients with an mRS score of 0-4 were included who had family caregivers.^15^ We excluded patients who had mRS scores of > 4, were mentally unstable and/or cognitively impaired (diagnosed cases), participated in other clinical trials, and patients with multi-organ failure or terminal stage.

### Randomization

We used a computer-generated stratified randomization technique considering participants age <65 years and ≥65 years based on studies carried out in China and got a sequence of numbers for which allocation participants would be assigned (intervention or control).^16, 17^ As the statistics for the age of patients with recurrent stroke and the types of strokes (hemorrhagic stroke, large artery atherosclerosis, cardioembolic, small vessel occlusion, lacunar, transient ischemic attack) were not available at the study site, we used that study findings as a reference, as recurrence rates varied depending on the age and type of the stroke.^18^ We used a sealed envelope containing the serial number with the allocated randomization group (either intervention or control). This randomization procedure was done by a research person (a head nurse of the ward) who was not involved in this study intervention or analysis.

### Sampling Technique and Study Procedures

A convenient sampling technique was used to enroll the participants. Neurologists of the hospital and Research Assistants (RA) nurses identified the participants from the inpatient departments before discharge, outpatient stroke clinic, and emergency department at NINS&H. The RA nurses explained the purpose of the study and obtained written informed consent from the participants. In case, when the participants were unable to take care of themselves and needed family caregivers’ support for their daily activities, we took consent from the family caregivers instead of patients considering them as participants. Then the RA nurses checked the eligibility criteria by referring to the medical record and enrolled the participants in this study. We allocated the participant’s randomization group by opening the sealed envelope. RA nurses obtained the participant’s sociodemographic information including age, gender, marital status, education, occupation, medical history, comorbidities, treatments, prescribed medications, types of stroke, mRS (0-4), risk factors of stroke, living status, clinical and laboratory data including glycated hemoglobin (HbA1c), Lipid profile, blood pressure (BP) by interviewing them and from the hospital medical records.

### For the intervention group (IG)

After enrollment at the hospital, at baseline RA nurses conducted a face-to-face health education and video presentation to the patients and their family caregivers for a total of 45 minutes. Then, RA nurses follow up the participants through phone calls every month (1st month to 3rd month: twice a month, and 4th month to 6 months: once a month) and send short messages (SMS) for recording their daily health education book. At midline after 6 months, RA nurses called the participants to attend the hospital for their follow-up visits, provided the same health education as baseline, collected data, and did laboratory tests.

**Figure 1.**
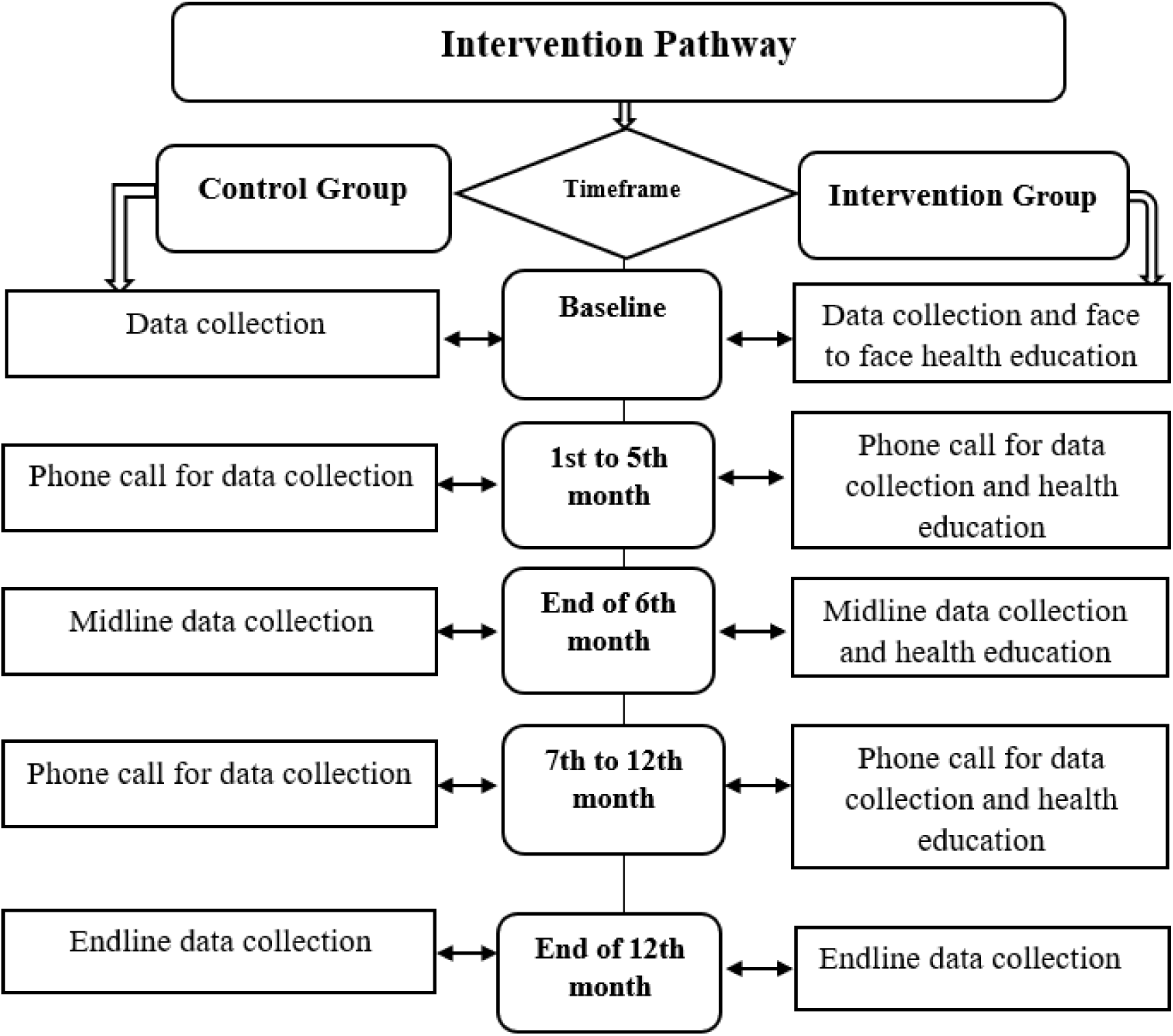
Study activities of intervention and control group.

Then, RA nurses follow-up the participants through phone calls monthly for 7 to 12 months and send the same short messages (SMS). After 12 months at the endline, RA nurses again called the participants to visit the hospital for their follow-up visit including data collection and laboratory testing. If the patient and family caregiver were unable to visit the hospital for any reason for the 6th month and 12-month follow-up visits at the hospital, then RA nurses provided health education over the telephone and advised to do the laboratory test from the nearby hospitals. RA nurses also visited the participants’ home for health education and sample collection if needed. If an RA nurse failed to communicate with the patient/caregiver over a phone call for 3 consecutive months considered as a dropout. This study activities were described elsewhere.^19^

### Intervention

The health education booklet contains the definition of stroke, its types, symptoms, and preventive strategies. It included the factors that influence the occurrence of stroke and its control like physical exercise, maintaining a healthy weight, maintaining healthy cholesterol, quitting smoking/smokeless tobacco, reducing intake of alcohol, managing stress, regular intake of medicine, having a balanced diet regularly, avoid junk/fatty foods. It was mentioned how to properly measure and intake salt, BP, and take medication. There was a provision to record all the daily measurements and activities of BP, exercise, and medication intake in the booklet at their home. For the measurement at home, we provided digital BP machines, medication boxes, and salt-measurement spoons. For visualization and easy understanding, we prepared and showed a video having the same health education content.

### For the control group (CG)

The CG participants received a one-time phone call from RA nurses every month to keep in contact with the study team (phone calls did not include health education) and received usual care from the hospital to which they were entitled. However, all the participants received a recording notebook (“stroke-friendly booklet”) at the end of the study to reduce disparities. On request, participants also received a brief health education based on the recorded notebook.

### Sample Size

We considered stroke recurrence rate was 20%^8^ and the incidence rate of recurrence was 1.9/100 and 3.8/100 person-year for the intervention and control group, respectively, 50% reduction^12^ and 80% power. The expected proportion of outcomes from the intervention group was 10%. Where, n =sample size, according to assumption P_1_= proportion of outcome from intervention group= 0.10 (10%), P_2_ = proportion of outcome from control group= 0.20 (20%), α= level of significance =0.05 (5%), 1-β= power of test=0.80 (80%), Z_1-α/2_= Z alpha value level of significance (1.96), Z1-β= Z beta value level of corresponding level of power (0.84). The estimated sample size was 196 for each arm (total 196X2=392 for both arms). With consideration of 10% dropout, the final sample size was 432 (216 for each arm).

### The operational definition of the terms for this study Recurrence of Stroke

Defined as a new stroke event occurring at least 28 days after the incident event,^16^ confirmed by a physician who treats and diagnoses the patients in this study. We considered this definition of recurrence >28 days to see the intervention effects to reduce stroke recurrence. However, different studies defined the recurrence of stroke as >24 hours after the onset of the incident stroke.^20^

### Adverse Events

Adverse event, including all causes of death and cardiovascular events, refers to any case that requires a diagnosis. For instance, a doctor treating and diagnosing the study participants confirmed any recurrence of stroke, new onset of acute myocardial infarction and unstable angina, heart failure, and peripheral arterial disease required hospitalization.

### Outcomes

The primary outcome of this study was to reduce the recurrence rate of stroke by providing health education after 12 months of follow-up. The secondary outcomes were to reduce (1) all adverse events (stroke recurrence, all causes of death, and cardiovascular events) and (2) increase medication adherence by providing health education after 12 months of a follow-up period.

### Data Monitoring and Quality Assurance

We conducted pre-testing of the questionnaire among 5% of the estimated samples (432) to understand the study procedures. Those patients were not included in the main study. In our study, investigators (medical professionals) had direct involvement in patient care. The investigators hold regular biweekly meetings with the RA nurses to discuss the progress of the study and resolve any confusion and concern. Furthermore, as part of routine quality control checking, investigators did field testing by independently verifying 5% of the collected data from study participants on the same day. The investigators observed the RA nurses’ consent-taking procedure, and data collection and performed physical examinations. At the field location, mistakes were fixed right away. RA nurses consulted with the investigators for any questions. The questionnaires were completed by RA nurses, who then input the information into a password-protected computer. Investigators examine the information and answer the questions.

### Statistical Analysis

We conducted an intention-to-treat (ITT) analysis for our study. Descriptive statistics were done to assess the baseline data. Categorical variables were reported as frequency and percentages and compared between the IG and CG by the chi-square (*X*^2^) test. Continuous variables were reported as mean and standard deviation, and after checking the normality test, we performed the Mann-Whitney U test as our variables were not normally distributed. To estimate the time to events for the 12-month follow-up period from enrollment to the occurrence of events, study completion, dropout, and death, we used Kaplan-Meier estimates and compared them between the IG and CG using the log-rank test. To compare the primary and secondary endpoints between the IG and CG, we used Cox-proportional hazards regression and results were presented as hazard ratios (HR) with a 95% confidence interval. Because statistical significance was observed in mRS at baseline, we adjusted for mRS by including it as a covariate in the Cox regression model to mitigate its potential impact on the primary and secondary outcomes. We also conducted chi-square test for medication adherence to compare the two groups. The significance level was set as p <0.05. Data was analyzed using SPSS (version 27.0; IBM Corp).

## RESULTS

Of the 760 participants who met the inclusion criteria and after excluding 328 participants, 432 consented to participate in the study. Consequently, participants were equally allocated into the intervention group (IG) (n=216) and the control group (CG) (n=216). After 12 months of follow-up, 307 (71.1%) participants completed the study; 166 (76.9 %) from the IG and 141 (65.3%) from the CG. The total dropouts were 30 (6.9%); 11 (5.1%) in the IG and 19 (8.8%) in the CG (Figure 2).

**Figure 2.**
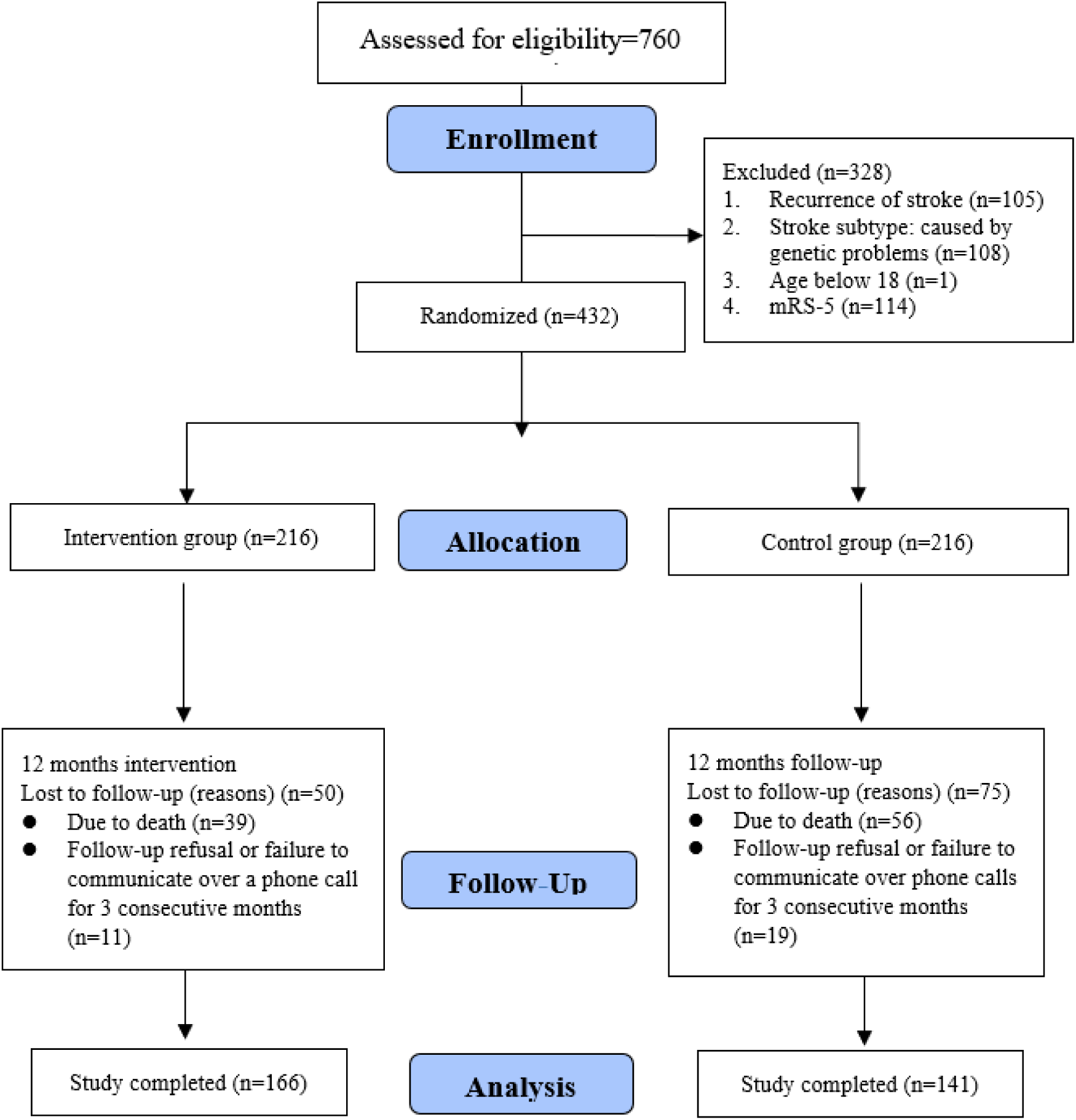
CONSORT study flow chart.

### Sociodemographic characteristics and clinical profiles of participants

The average age of the participants was 55.2±13.5, male 273 (63.2%) and 169 (39.1%) from the urban areas. The mean duration of onset of stroke to enrollment (*P*<0.001), dyslipidemia (*P=*0.03), taking hyperlipidemic medication (*P=*0.049), and mRS score 1, 2, and 3 (*P*<0.001) were significantly higher in the IG compared to CG. In contrast, compared to IG, CG had significantly more urban residence (*P*=0.023), history of myocardial infarction (*P=*0.008), and mRS score 4 (*P*<0.001). Age and stroke types were well allocated, however, mRS was statistically significantly higher in the CG (Table 1).

**Table 1.**
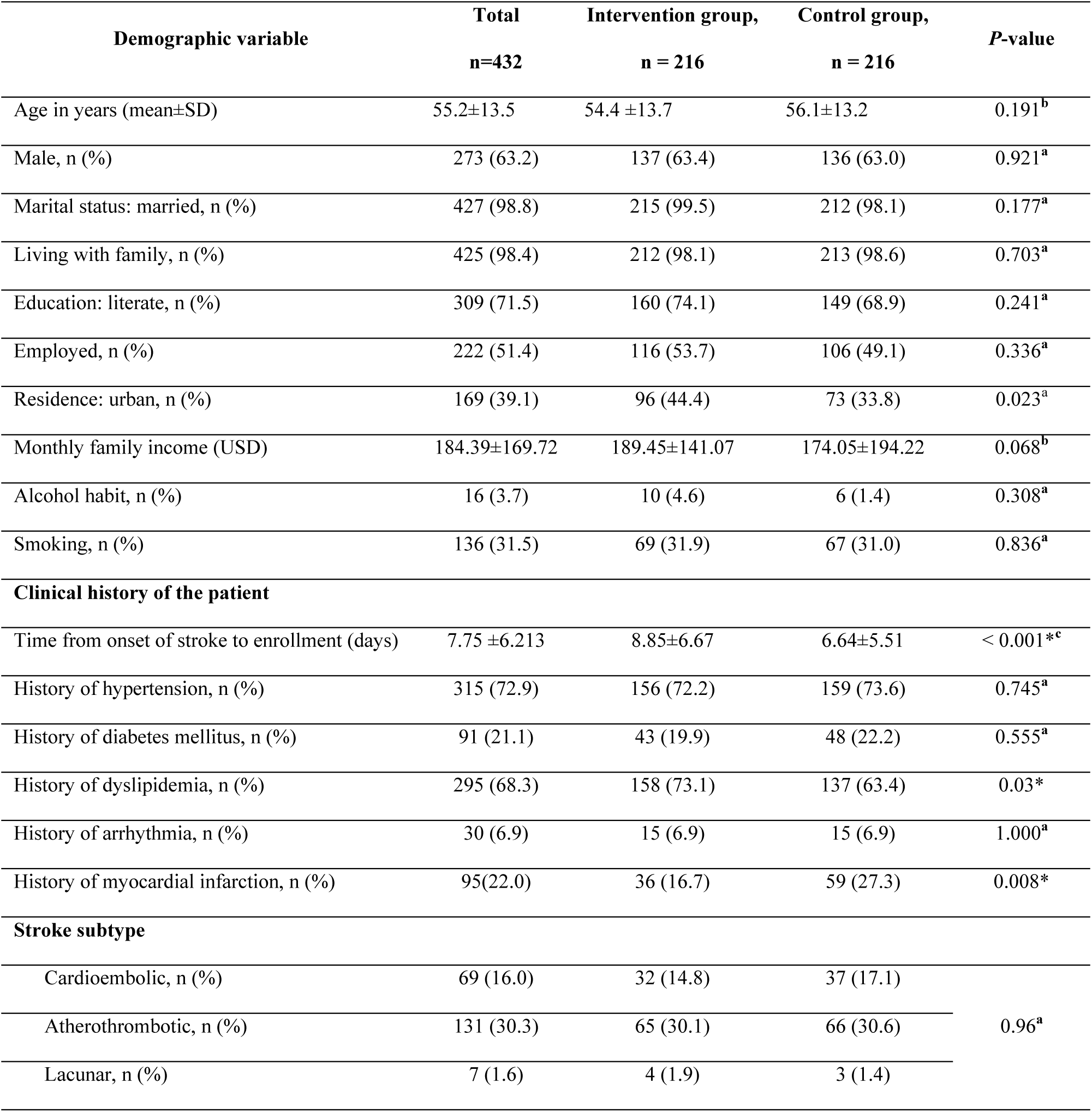
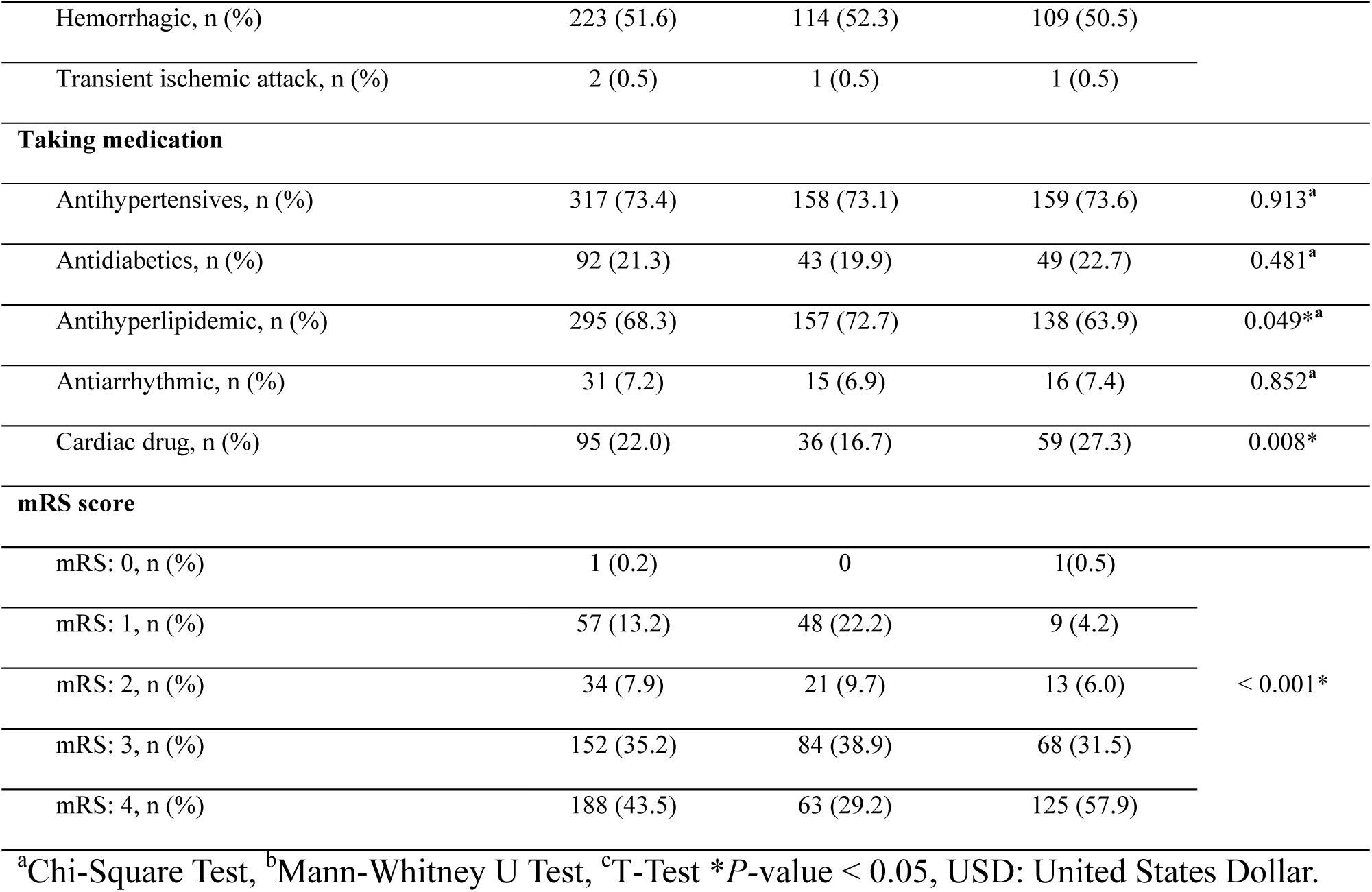
Sociodemographic characteristics and clinical profiles of participants.

### Primary endpoint

The monthly distribution of recurrent stroke is shown in Supplementary Table 1. We found that the first month had 27 recurrences (56.3% of total recurrences and 11.1% of total participants) occurred; within the first 6 months, 40 recurrences (83.3% of total recurrences) and remaining recurrences occurred in the next 6-month follow-up (n=8).

The primary endpoint of this study was the recurrence of stroke after 28 days to 12 months which was 22 (5.1%) (14 (6.5%) in the IG and 8 (3.7%) in the CG); the difference was not significant (*P*=0.189) between the groups (Table 2).

**Table 2.** 12-month follow-up among recurrence.

| Recurrence | Total n=432<br>(%) | Intervention group<br>n = 216 (%) | Control group<br>n = 216 (%) | *X*<sup>2</sup> value | *P*-value |
| --- | --- | --- | --- | --- | --- |
| After 28 days | 22 (5.1) | 14 (6.5) | 8 (3.7) | 1.724 | 0.189 |
| within 28 days | 26 (6.0) | 11 (5.1) | 15 (6.9) | 0.655 | 0.418 |
| Total | 48 (11.1) _ | 25 (11.5) | 23 (10.6) | 0.094 | 0.759 |

Chi-Square Test

### Secondary endpoints

The total recurrence of stroke was 48 (11.1% of total participants); among them 15 (3.5%) were alive and 33 (7.6%) died. The secondary endpoints were the stroke-related all adverse events including recurrence, death, and cardiac events. All events were 116 (26.9%); whereas all events were 54 (25.0%) and 62 (28.7%) in the IG and CG respectively, and the difference was not significant [Hazard ratio (95% CI): 1.216 (0.844-1.752); P=0.294]. Total death accounted for 95 (22.0%); 39 (18.1%) in the IG and 56 (25.9%) in the CG, and the difference was significant [Hazard ratio (95% CI): 1.531 (1.017-2.304); *P*= 0.041]. However, after adjusting the covariate (mRS of baseline), the deaths were found not significant [Hazard ratio (95% CI): 0.818 (0.540-1.238); P = 0.341]. The monthly distribution of all causes of death was calculated and shown in Supplementary Table 1. A half (n=47, 49.5%) of deaths occurred within the first month. Total cardiac events were 8 (1.8%), 5 (2.3%) in the IG, and 3 (1.4%) in the CG (Table 3). We also saw the distribution of different mRS groups (0-2), 3 and 4, and found mRS 4 had more deaths followed by mRS 3 and 0-2 (Figure 3).

**Table 3.**
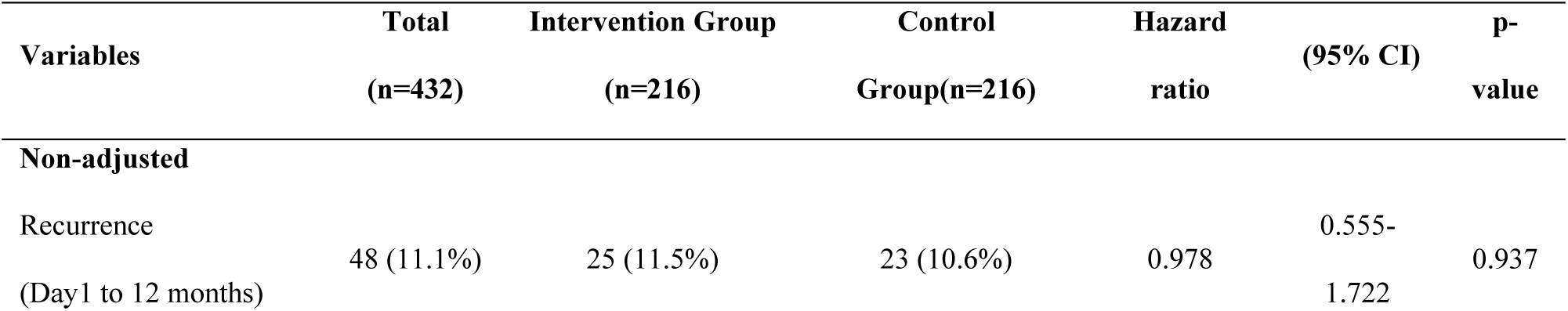
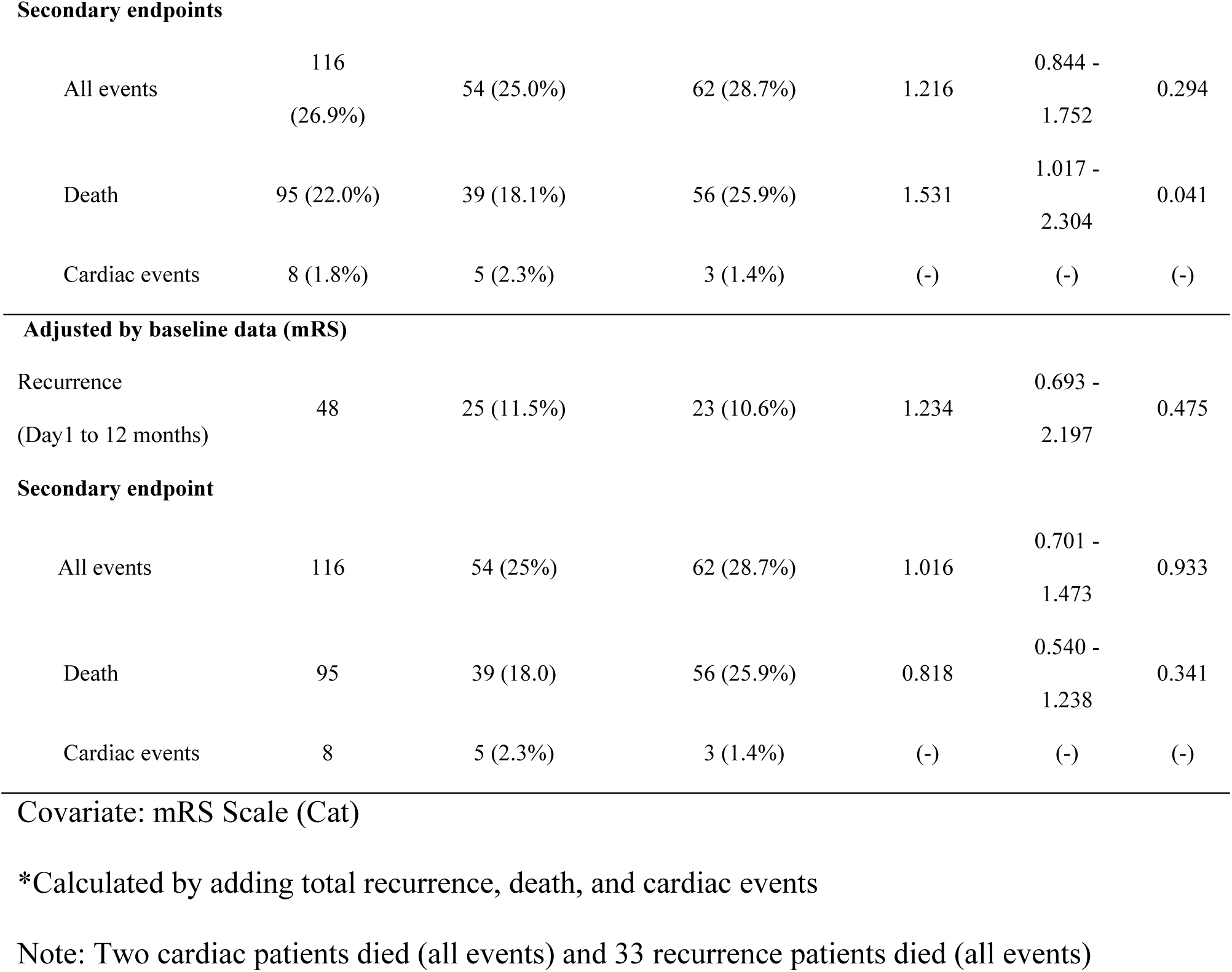
Cox-regression of the endpoints.

**Figure 3.**
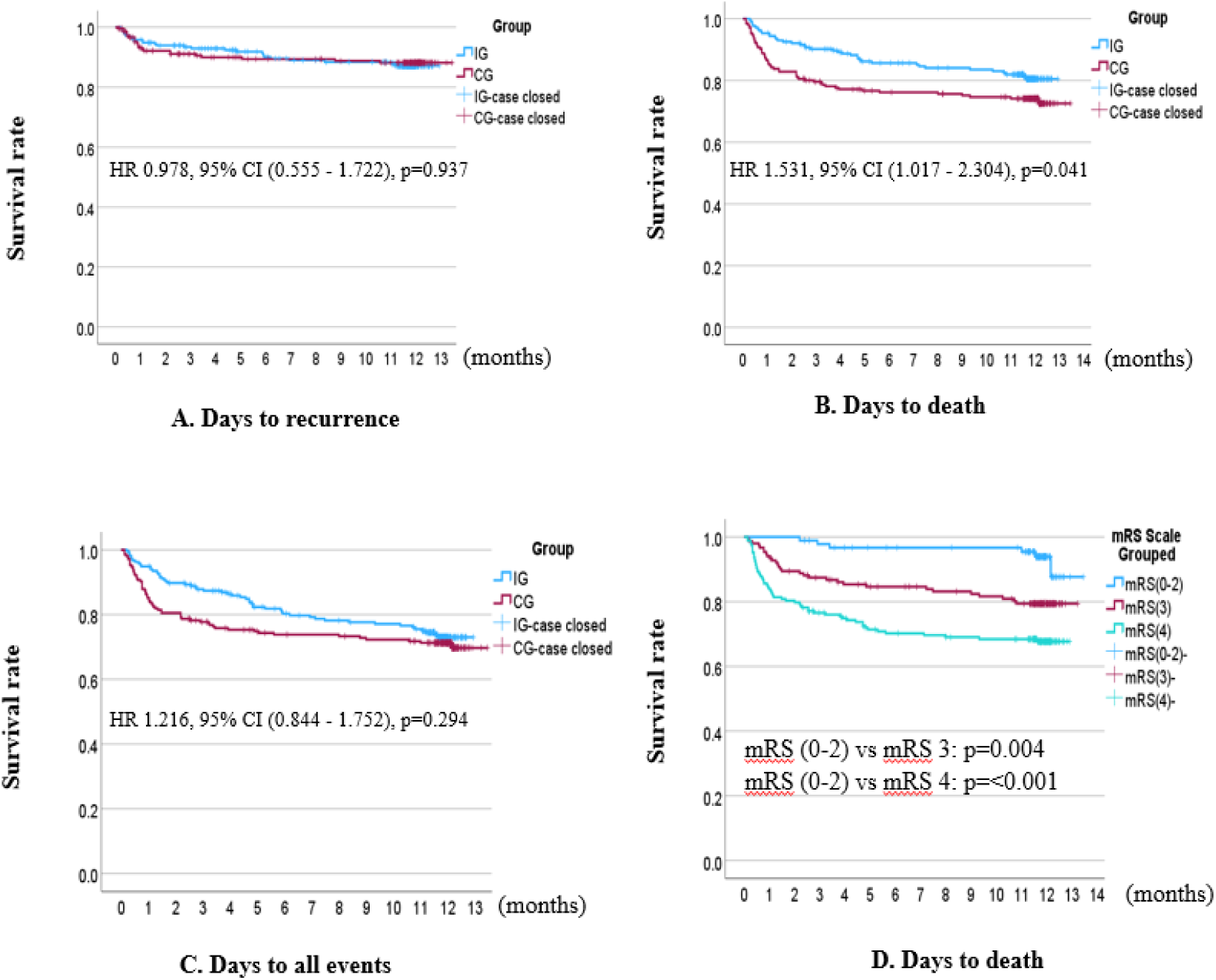
Survival rate during 12 months from enrollment. A, Recurrence; B, Death; C, All events; D, mRS scale (0-4). IG, Intervention group; CG, Control group; HR, hazard ratio.

### Kaplan Meier Graph

Kaplan-Meier survival curve showed that the estimated survival days for 12 months follow-up for mRS (0-2) was 401.8 days [95% CI: 388.7-414.9], mRS (3) 348.4 days [95% CI: 327.4-369.4], and mRS (4) 293.5 days [95% CI: 270.3-316.6]. The death was found significantly higher in mRS (3) (p=0.004) and mRS (4) (p<0.001) compared to mRS (0-2) (Figure 3D).

We calculated the recurrence and death by type and mRS within 12-month follow-up period and found the recurrence and death were the highest in the hemorrhagic (23 (47.9%), 48 (50.5%)) and mRS 4, 31 (64.6%) and 59 (62.1%), respectively (Table 4).

**Table 4.** Distribution of recurrence and death of stroke by types and mRS within 12-month.

| Types of stroke | Recurrence (n=48) | Death (n=95) |
| --- | --- | --- |
| Cardioembolic | 11(22.9%) | 26 (27.4%) |
| Atherothrombotic | 14 (29.2%) | 20 (21.1%) |
| Lacunar | 0 | 1 (1.1%) |
| Hemorrhagic | 23 (47.9%) | 48 (50.5%) |
| Transient ischemic attack | 0 | 0 |
| **mRS** |  |  |
| mRS 0 | 0 | 0 |
| mRS 1 | 3 (6.3%) | 2 (2.1%) |
| mRS 2 | 2 (4.2%) | 4 (4.2%) |
| mRS 3 | 12 (25.0%) | 30 (31.6%) |
| mRS 4 | 31 (64.6%) | 59 (62.1%) |

**Supplementary table 1.**

Medication adherence strongly affects the recurrence of stroke, we compared the medication intake adherence between the IG and CG at baseline, midline, and endline. We found significant improvement in the IG at midline and endline (p<0.001 and p=0.002, respectively). Taking medication “as doctor’s prescription for 6-7 days per week” was significantly increased (Table 5).

**Table 5.** Medication Adherence.

| Groups | Not at all | Occasionally | Once/week | 2-3 days /week | 4-5 days/week | 6-7days/week | P-value |
| --- | --- | --- | --- | --- | --- | --- | --- |
| Baseline | n (%) | n (%) | n (%) | n (%) | n (%) | n (%) |  |
| IG (n=216) | 67 (31.0) | 54 (25.0) | 1 (0.5) | 6 (2.8) | 23 (10.6) | 65 (30.1) | 0.276 |
| CG (n=216) | 57 (26.4) | 71 (32.9) | 0 (0.0) | 2<br>(0.9) | 25 (11.6) | 61 (28.2) |  |
| Midline* |  |  |  |  |  |  |  |
| IG (n=149) | 12 (8.1) | 2 (1.3) | 0 (0.0) | 10 (6.7) | 4 (2.7) | 121 (81.2) | <0.001 |
| CG (n=130) | 26 (20.0) | 1 (0.8) | 0 (0.0) | 0 (0.0) | 0 (0.0) | 103 (79.2) |  |
| Endline |  |  |  |  |  |  |  |
| IG (n=166) | 2 (1.2) | 0 (0.0) | 7 (4.2) | 5 (3.0) | 11 (6.6) | 141 (84.9) | 0.002 |
| CG (n=141) | 11 (7.8) | 0 (0.0) | 18 (12.8) | 5 (3.5) | 6 (4.3) | 101 (71.6) |  |

Chi-Square Test, n= number, %= percentage,
\* Patient attendance was low at midline

## DISCUSSION

This is the first intervention study that followed stroke patients for 12 months to evaluate the risk factors for recurrence and other adverse events related to stroke in Bangladesh. This study findings can be used as the most reliable evidence for epidemiological figures in Bangladesh. We provided health education for controlling the modifiable risk factors to prevent the recurrence of stroke who had stroke attack.

We compared the patients who received health education intervention with those who received only usual care and found that there was no significant difference in reducing the recurrence of stroke between the IG and CG after 12 months of follow-up. A similar finding was observed in another study that recurrence could not seem to be reduced by only self-management education with treatment coordination.^12^ A hospital-based 12-month survey was conducted in Bangladesh (one hospital, n=150) to estimate the stroke recurrence rate and observed that without any intervention the recurrence occurred in 20% of the stroke patients. In our study after the intervention, we found the recurrence rate was 11.1% within the same period (day 1 to 12 months), which may reflect our intervention works.^8^ In Japan, one meta-analysis discovered that the recurrence rate was 7.2% in a 3.9-year follow-up study. ^21^

We found that more than half of the recurrence occurred in the first month after the first stroke attack. A few of them were diagnosed as recurrence of stroke one day after enrollment. Therefore, it is very difficult to evaluate the effects of intervention within this short time. Many patients were first diagnosed with new comorbidities such as hypertension, diabetes and dyslipidemia after attending the hospital and started treatment. Therefore, the risk of recurrent stroke remained after stroke and made the patients more vulnerable to other adverse events. It also increased the risk of attack after discharge from the hospital for inadequate monitoring of the risk factors at home. Though the participants were on antihyperlipidemic medication for their dyslipidemia, there was still a high risk of developing stroke recurrence.^22^ These might act as confounding factors which does not reduce significantly the recurrence of stroke in the IG.

We found within the first 6 months, the recurrence rate was slightly decreased in the IG compared to the CG. However, after six months to twelve months, the recurrence rate increased in the IG, even continuing the same intervention. The risk of recurrence in our study was higher within the first six months compared to the next six months in our 12-month follow-up period. Another study also found that the first three months are crucial for the recurrence of stroke.^8^ Therefore, if we can control the risk factors within this time, we can prevent the recurrence mostly.

As recommended for the diagnostic procedure of stroke,^23^ after visiting healthcare facilities, physicians performed laboratory investigations such as computerized tomography (CT) scan and magnetic resonance imaging (MRI). However, most of the patients in this study could not perform these tests for their subsequent attack to diagnose recurrence and other adverse events including the cause of death due to unavailability of investigation facilities and patient’s financial constraints. Therefore, it could not be possible to diagnose the cause of illness of the stroke patientsIn addition, many of our patients could not perform these tests as they died at home before attending the hospital for their diagnosis; those deaths were considered as unknown causes. Among the recurrent patients, less than three-quarters (33/48) of them died. Therefore, recurrence was considered as a main risk factor for death.^24^ We estimated the death of the participants and found that overall death was significantly reduced in the IG group compared to CG during the 12-month follow-up period. However, after adjusting mRS at baseline as a covariate, death was found not significant. One study also concluded the same way that health education had a positive impact on reducing post-stroke death.^25^ We identified the cause of death mainly as the recurrence of stroke followed by cardiac problems, liver chlorosis, cancer, and other complications. Out of 8 cardiac events, 3 and 5 died in the CG and IG, respectively and cardiac events were not significantly reduced after intervention.

We found mRS 4 (the most severe condition of the participants) was significantly higher in the CG compared to the IG group and it might cause significantly higher deaths in the CG. As most (78.7%) of our patients had mRS 3 and 4, therefore, we experienced an extremely higher death (22%) in this study population compared to another study conducted in Japan (10.3%) with a 14-year follow-up.^26^ We recruited mRS 0-2 disproportionately compared to mRS 3-4 in both IC and CG, this might be influenced the effects of intervention benefits. In Bangladesh, patients with stroke attacks usually attend the local healthcare facilities. As there are weak primary and secondary healthcare systems, patients are not well diagnosed and treated (lack of qualified physicians and diagnostic facilities) and miss the opportunity to receive acute care management. Therefore, patients who were referred to this tertiary hospital without receiving any standard acute treatment such as tissue plasminogen activator (tPA) and/or surgical intervention during the golden time window makes the patients more vulnerable to develop recurrence and other adverse events.

Moreover, there is no national health insurance system exist in Bangladesh,^27,28^ most of the treatment cost of the patients was incurred out of their pocket. Stroke patients with mRS 0-2 rarely visit the hospital; only in severe cases, patients are referred and attended to the hospital. NINS&H is the only specialized referral hospital in Bangladesh and receives more patients compared to its limited beds. Therefore, only severe stroke patients (mRS 3-4) attended and were admitted to the hospital. To get stroke patients on different scales (0-2), with the inpatient department, we also enrolled participants from the outpatient department to include patients with mRS 0-2.

The average age of our participants was 55 years with a minimum of 18 to a maximum of 92 years and males were predominant. One meta-analysis conducted in Japan reported that the average age of the patients was 67.2 years.^21^ It is alarming that young people are also at risk of stroke attacks in this population. Some of them were vulnerable to stroke as they had modifiable risk factors like smoking.^29,30^ Three-quarters of them had a history of hypertension, and less than three-quarters had dyslipidemia, those are the major risk factors for the recurrence of stroke.^22,31^ There is no health checkup system in Bangladesh for all, and most of the younger population is not aware of their health status and are not treated properly. This study strongly indicated that campaigning and introducing regular health checkup systems should be incorporated in the national health policy. This study observed participants’ medication adherence was significantly improved during their monthly follow-up in the IG compared to CG. However, medication adherence also improved in the CG throughout the study period, which might be due to providing regular phone calls without any intervention.

The sociodemographic attributes of the participants may also be a factor in determining the present results. About half of our participants were unemployed, and some of them had financial constraints. Our participants’ average income was far below (66.6% of the participants) than the national average income of the individuals (US dollars 277).^32^ Therefore, they had limited access to hospitals or any healthcare facilities for their regular health checkups and laboratory examinations. Moreover, considering their age, elderly individuals feel inconvenienced when attending healthcare facilities due to the long distance from their homes. Despite the desire and necessity of many participants to attend the hospitals, financial constraints prevented them from covering the treatment-related associated expenses. Without national health insurance coverage and any financial support, it is very difficult to expand costly treatment costs from their own pocket. It made more participants withdraw from the study.

### Strengths

In our study, we provided health education to stroke patients and their family caregivers and trained them on how to measure BP, salt, and regular intake of medication first in Bangladesh. We supplied health education booklet, BP machine, salt measuring spoon, and medication box to use at the participants’ home to make them empowered. Moreover, we conducted our study in the only tertiary national specialized neuroscience hospital, the study findings can be generalized for the patients of Bangladesh and other countries. It was the comparatively large sample size

### Limitations

The study has certain limitations. We faced potential biases due to heterogenicity in baseline illness levels and consciousness among different subtypes of stroke patients, and the need for patient and caregiver education on self-management. During the enrollment, we did not consider the severity of the patients (mRS), usually stroke subtype and mRS are correlated,^33^ Therefore, there was an uneven distribution of the participants, which influenced the findings of our study. A huge number of participants died at their homes without any investigation for the diagnosis of cause of death. These limited diagnostic facilities, lack healthcare resources and economic difficulties of patients influenced our study findings.

## CONCLUSIONS

We provided health education to our study participants and found recurrence of stroke and all other adverse events were not decreased. The participants experienced more recurrences and deaths in both groups in their first month after stroke compared to the remaining eleven months and death was found significantly less in the first month in the IG compared to the CG. Overall hemorrhagic stroke and mRS 4 had more recurrences and deaths in both groups. Therefore, the acute care system needs to pay more attention to prevent recurrences and death. We could not identify the cause of death and recurrences of the participants who had not attended the healthcare facilities for their diagnosis and treatment. This study emphasizes the importance of health education in enhancing medication adherence among patients. It suggests that comprehensive health education could improve treatment outcomes and foster long-term behavioral changes, thus enhancing patient well-being and healthcare effectiveness. This study findings may help to assist further development of future health interventions inclusively all participants for the prevention of the onset recurrence of stroke. A sustainable strategy needs to be developed for effective stroke-preventive strategies in the public domain. Further interventional study with a large sample size by addressing all limitations including diagnostic facilities for stroke will determine the influencing factors for the development of recurrence of stroke and death.

## Data Availability

The data supporting the findings of this study are available from the corresponding author upon reasonable request.

## Acknowledgments

We would like to express our sincere gratitude to the study participants. We extend our thanks to the NINS&H authority, for their support and the Research Assistant nurses and laboratory staff for their unwavering assistance mainly: Israt Jahan Eti, Naznin Nahar, Shorifunnahar, Shika Parven, Tarana Manju Joya, Salma Akter, Sipra Mondal, Shakil Ahmed, Md. Kayes Azad Polok, Raisa Islam, Mostarina Shopna, Pronoy Bonik, and Suraiya Akter, Gita Roy.

## Source of Funding

The study was supported by the research grant from Hiroshima University, Japan

## Disclosures

None reported.

## Notes

### Competing Interest Statement

The authors have declared no competing interest.

### Clinical Trial

NCT05520034

### Author Declarations

This study was approved by the Institutional Review Board and the Ethical Review Board of the NINSH (IRB/ NINSH/2022/151).

